# Maternal and cord-blood inflammatory markers and BDNF in diabetic vs non-diabetic pregnancies

**DOI:** 10.1101/2025.07.02.25330741

**Authors:** Michael P. Wendel, Heather L. Moody, Eric R. Siegel, Hari Eswaran, Shannon Rose

## Abstract

**Background:** Maternal diabetes is associated with increased systemic inflammation and has been linked to adverse neonatal outcomes, including developmental delays that persist into early childhood. In this study we sought to characterize and compare the maternal levels and fetal cord-blood levels of the inflammatory markers C-reactive protein (CRP) and IL-6, as well as the neurotrophin brain-derived neurotrophic factor (BDNF) between mothers with pre-gestational Type-1 diabetes (T1DM) or Type-2 diabetes (T2DM), and non-diabetic controls (nonDM).

**Methods:** A prospective cohort design was employed, analyzing biomarker concentrations during the third trimester in 98 pregnant women ages 18-40 years of age including 16 participants with T1DM, 49 participants with T2DM and 33 control participants matched for gestational age and body mass index (BMI) to control for confounding factors such as obesity. Plasma samples were collected at 28–30 weeks, 34–36 weeks, delivery, and from cord blood. The biomarkers CRP, IL-6, and BDNF were measured using standardized assays, and concentrations were compared among groups using including ANOVA.

**Results:** In T2DM mothers, CRP levels were 2x higher in the third trimester as compared to nonDM controls. In T1DM mothers, IL6 levels were 3x lower than nonDM controls and 3.4x lower than T2DM. While not reaching statistical significance, cord-blood levels of IL6 were higher in T2DMs than other groups (p=0.052). When examining BDNF levels, no differences were observed between groups.

**Conclusions:** This study emphasizes the importance of addressing inflammation-related risks in pregnancies affected by diabetes. Targeted interventions may mitigate adverse neonatal outcomes and improve health trajectories. Future research should explore direct pathways linking maternal inflammation to fetal neural function to inform clinical strategies.

## 1. Introduction

### Background

Pregnancies affected by maternal diabetes are slowly increasing in prevalence. As such, about 1-2% of all pregnancies are now complicated by pregestational diabetes.^1^ Type 1 diabetes is characterized by an autoimmune destruction of β cells in the pancreas, leading to a deficiency in insulin production. Typically, this occurs in younger patients. Type 2 diabetes, on the other hand, is typically associated with peripheral insulin resistance. This condition is more commonly diagnosed later than type 1 diabetes and is associated with obesity. The current obesity epidemic has led to type 2 diabetes being the predominant form of pregestational diabetes in the United States.^1^ Bardenheimer, et al. found that deliveries to pregestational diabetic mothers has increased across the United States. In 2000, there were about 13,000 deliveries complicated by pregestational diabetes across 19 different states. In 2010, that number had increased to over 18,000, which translates to a 38% increase in maternal diabetes in a 10-year time span. In all 19 states, the increase in deliveries complicated by pregestational diabetes was statistically significant. In addition, the increase was significant in all age groups and ethnicities as well.^2^ As the number of patients with diabetes increases, this will become a much more pervasive factor complicating pregnancies.

Diabetes has a number of adverse effects on pregnancy and the developing fetus. Patients with pregestational diabetes (and particularly those with poorly controlled diabetes) are at risk for congenital malformations, fetal and neonatal growth disorders, stillbirth, perinatal death, increased need for cesarean section, and preterm delivery among many other poor outcomes.^3^ Of particular interest to this study, there seems to be an effect on neurodevelopmental outcomes in the offspring of diabetic mothers. As early as 1969, for instance, Churchill, et al. found IQ scores of children born to diabetic mothers to be lower than those of children born to non-diabetic mothers.^4^ Frasier found a similar finding in a population study out of Sweden.^5^ Stehbens, et al. and Petersen, et al. found differences in cognitive abilities and language development in these children.^6-8^ Kimmerle, et al. found a general retardation in development of 90 children born to diabetic mothers.^9^ Hod, et al. found neurodevelopmental delays in the first year of life.^10^ Ornoy, et al. and Rizzo et al. found normal cognitive function, but delays in motor development in this population.^11,12^ In addition to delayed neurodevelopment, some studies have suggested maternal diabetes is linked to diagnoses like attention deficit-hyperactivity disorder and autism spectrum disorders.^13-18^

This then begs the question, what property of pregnancies complicated by diabetes causes abnormal development of the fetal brain. A number of recent studies have focused on the effects of inflammation and the relationship between higher levels of inflammatory markers or inflammatory effects on circulating neurotrophins as they relate to fetal neural development. Maternal inflammatory-pathway activation may have effects on the developing fetal brain and subsequent behaviors.^19^ For instance, increased levels of the inflammatory cytokine

IL-6 may be a marker for neonatal neuronal damage.^20^ IL-6 elevations are also associated with effects on working memory even two years after birth.^21^ Elevations of another inflammatory marker, C-reactive protein (CRP), have been associated with schizophrenia diagnosis in offspring.^22^ CRP has also been implicated in fetal growth restriction and small for gestational age neonates.^23^ Restriction of overall fetal growth shows that maternal inflammation can have overall deleterious effects on fetal development which may also impact the fetal brain. Studies thus far have not been able to determine if these are direct effects on fetal development or if elevated CRP is a marker for abnormal development.

A third factor of recent research has been how the biomarker brain-derived neurotrophic factor (BDNF) is related to fetal neural development. BDNF has been shown to promote neuronal growth, morphology, and function as well as maintaining survival of those neurons by inhibiting apoptosis.^24-26^ In adults, low levels of BNDF have been seen in neurodegenerative diseases such as Alzheimer’s disease.^25,26^ It may be the primary neurotrophin involved in fetal brain development.^24^ Mouse models have shown that at least a portion of fetal BDNF is obtained via placental transport, so if maternal BDNF production is altered, it could affect fetal brain development.^27^ Krabbe, et al. found that patients with type 2 diabetes have lower levels of BDNF in the serum and produce less BDNF in response to high levels of blood glucose.^28^

The long-term goal of our ongoing study is to determine whether maternal levels of CRP, IL-6, and BDNF during the 3^rd^ trimester of pregnancy mediate some of the adverse effects of pregestational diabetes on fetal and postnatal neurodevelopment. The immediate objective of this report is to characterize and compare the maternal levels and fetal cord-blood levels of CRP, IL-6, and BDNF between Type-1 diabetics, Type-2 diabetics, and non-diabetic controls.

## 2. Methods

### 2.1 Subjects

This study was approved by the University of Arkansas for Medical Science’s Institutional Review Board, and all participants provided informed consent to participate in the study. The inclusion criteria were simply patients aged 18-40 with preexisting diabetes or diabetes diagnosed in the first trimester. A low-risk cohort was also recruited prospectively by matching non-diabetic subjects by BMI class (BMI ≥30 kg/m^2^ versus not) and gestational age to diabetic subjects. Exclusion criteria included maternal ages less than 18 or over 40, fetal malformations, fetal genetic anomalies, and non-singleton pregnancies. Table 1 provides the overall demographics of the participants. There were 98 patients recruited: 33 non-diabetic patients, 16 type 1 diabetic patients, and 49 type 2 diabetic patients. Patients were asked to present for blood draws at 3 separate points in pregnancy as described below. To encourage compliance with visits, patients were offered $80 for each event in which they participated.

**Table 1:** Participant Characteristics.

| **Table 1: Participant Characteristics** |  |  |  |  |
| --- | --- | --- | --- | --- |
| **Participant Characteristics** | **nonDM<sup>1</sup>**<br>**N=33** | **T1DM<sup>2</sup>**<br>**N=16** | **T2DM<sup>3</sup>**<br>**N=49** | **P<sup>†</sup>** |
| **Maternal Age, years** |  |  |  | 0.002 |
| Mean (SD) | 27.3 (5.0) | 26.7 (4.0) | 30.8 (4.8) |  |
| Minimum- -Maximum | 18- -37 | 20- -34 | 21- -41 |  |
| **Maternal Ancestry** |  |  |  | <.001<sup>‡</sup> |
| Hispanic, % (N) | 24% (8) | 0% 0 | 14% (7) |  |
| NH<sup>§</sup> Black, % (N) | 21% (7) | 13% (2) | 51% (25) |  |
| NH<sup>§</sup> Other, % (N) | 0% (0) | 6% (1) | 0% (0) |  |
| NH<sup>§</sup> White, % (N) | 55% (18) | 81% (13) | 35% (17) |  |
| **Pre-Pregnancy BMI, kg/m<sup>2</sup>** |  |  |  | <.001 |
| Mean (SD) | 32.8 (6.5) | 30.7 (6.5) | 39.0 (9.2) |  |
| Minimum- -Maximum | 23- -50 | 23- -44 | 24- -61 |  |
| **Gravidity** |  |  |  | 0.568 |
| 1, % (N) | 21% (7) | 19% (3) | 20% (10) |  |
| 2, % (N) | 21% (7) | 50% (8) | 31% (15) |  |
| 3, % (N) | 27% (9) | 12% (2) | 24% (12) |  |
| >3, % (N) | 30% (10) | 19% (3) | 24% (12) |  |
| *Mean Gravidity* | *2.94* | *2.50* | *2.78* |  |
| **Parity** |  |  |  | 0.433 |
| 0, % (N) | 27% (9) | 38% (6) | 33% (16) |  |
| 1, % (N) | 39% (13) | 44% (7) | 41% (20) |  |
| 2, % (N) | 12% (4) | 19% (3) | 12% (6) |  |
| >2, % (N) | 21% (7) | 0% (0) | 14% (7) |  |
| *Mean Parity* | *1.48* | *0.81* | *1.16* |  |
| **Hemoglobin A1c, %** |  |  |  | <.001 |
| Mean (SD) | 4.97 (0.82) | 6.76 (1.18) | 6.16 (1.30) |  |
| Minimum- -Maximum | 2.8- -6.7 | 5.4- -9.4 | 3.8- -10.5 |  |
| *Number non-missing* | *30* | *16* | *49* |  |

1:Non-diabetic group. 2:Type-1 Diabetes Mellitus group. 3:Type-2 Diabetes Mellitus group.
†P-values are from the Kruskal-Wallis test and ‡Fisher's exact test. §Non-Hispanic.

### 2.2 Sample collection

Approximately 8 ml of blood was collected from each mother at 3 separate points during pregnancy. The first sample was drawn at 28-30 weeks gestation (E1), the second at 34-36 weeks gestation (E2), and the last maternal sample was collected at the time of delivery (E3). Additionally, approximately 8 ml of venous blood was collected from the umbilical cord of her neonate immediately after delivery. Blood was collected into sodium heparin tubes and placed immediately on ice. Plasma was collected after centrifuging the blood for 15 min at 2000×*g* at 4 °C. Plasma was frozen and stored at - 80 ºC.

### 2.3 Analytical procedures

CRP was measured in both maternal and cord plasma (diluted 1:10,000-1:20,000 in assay buffer) by ELISA (Human C-Reactive ELISA, Cat# RAB0096, Millipore Sigma, Burlington, MA, USA)) following the manufacturer’s protocol. IL-6 was also measured in both maternal and cord plasma (diluted 1:2 in assay buffer) by ELISA (Human IL6 ELISA, Cat# RAB0306, Millipore Sigma) following the manufacturer’s instructions. BDNF was measured in maternal and cord blood plasma (diluted 1:6 in assay buffer) by ELISA (Human BDNF ELISA, Cat# RAB0026, Millipore Sigma) following manufacturer’s instructions. Samples were run in duplicate and data was acquired on a BioTek EPOCH2 microplate reader (Agilent, Santa Clara, CA, USA). Standard curves were plotted and results were extrapolated using Graph Pad Prism 10.

### 2.4 Statistical Analyses

The two prospectively enrolled control groups formed by matching on GA and BMI to each diabetes (DM) group were combined for analysis. There were three DM-status groups: Type-1 diabetics (T1DMs), Type-2 diabetics (T2DMs), and non-diabetics (NonDMs). Maternal ancestry was defined as consisting of 4 groups: Hispanic, Non-Hispanic (NH) Black, NH White, and NH Other. Maternal ancestry was summarized by DM status as number and percent in each ancestry group and tested for heterogeneity across status groups with Fisher’s exact test. Maternal age, pre-pregnancy BMI, and HbA1c at enrollment were summarized by DM status as mean, standard deviation (SD), and range, and tested for DM-status heterogeneity with the Kruskal-Wallis test. Maternal gravidity and parity, both being integer-valued scores, were summarized by DM status as the number and proportion at each score level and as a mean score, then tested for DM-status heterogeneity with the Kruskal-Wallis test. Sample concentrations of BDNF (pg/mL), IL-6 (pg/mL), and CRP (μg/mL) were transformed to their natural logs to stabilize variances and reduce right-skewing. The resulting log-values were summarized by DM status as means and SDs and tested for status-group differences with 1-way ANOVA (for BDNF and CRP) and unequal-variance ANOVA (for IL-6). Each ANOVA was followed by a single post-hoc comparison between T2DMs and nonDMs. Group differences in natural logs were exponentiated to yield the equivalent fold-changes in concentration. Analyses were performed with SAS v9.4 software (The SAS Institute, inc., Cary, NC, USA). All statistical tests employed an unadjusted P<0.05 significance level, despite the multiple comparisons, in order not to inflate Type II (false-negative) error.

## 3. Results

### 3.1 Patient Characteristics

There were 98 participants recruited: 33 non-diabetic controls, 16 type-1 diabetic patients, and 49 type-2 diabetics. These groups were analyzed for differences in age, ancestry, pre-pregnancy BMI, gravidity, parity, and initial-pregnancy hemoglobin A1c percentage. Groups were similar in gravidity and parity. On average, however, the type 2 diabetics were approximately 3 years older than controls and type 1 diabetics (*P*=0.002), with pre-pregnancy BMIs from 6.7 to 9.5 kg/m^2^ higher (*P*<0.001). These findings are not completely unexpected given that Type 2 diabetes diagnoses are more common in older patients with obesity.(27) Group differences in maternal ancestry were significant (*P*<0.001), but complex. Non-diabetic controls had the highest percentage of Hispanic patients (24%), type-2 diabetics had the highest percentage of Black patients (51%), and type-1 diabetics had the highest percentage of White patients (81%). Hemoglobin A1c percentages averaged roughly 5.0% in controls, 6.2% in type-2 diabetics and 6.8% in type-1 diabetics (*P*<0.001), as expected.

### 3.2 Plasma Markers Results

CRP, IL-6, and BDNF were measured in blood drawn at 28-30 weeks, 34-36 weeks, at the time of delivery, and from fetal cord blood. Some data points are missing due to early delivery, study protocol non-adherence, or missed appointments, which all lead to a different number of samples at each event time.

However, the number of missing data points is low overall with only at most 10 missing control data points, 3 missing type-1 data points, and 10 missing type-2 data points in any one collection during the study. In total, there were 252 maternal samples drawn across all groups in the 3rd trimester and there were 79 cord blood samples available for evaluation.

Maternal CRP concentrations differed among groups at 2 of 3 maternal time points (**Table 2**). When T2DMs were compared to nonDM controls, CRP levels were found to be 0.69 log units (1.99-fold or 1.99x) higher at 28-30 weeks (*P*=0.019), 0.77 log units (2.16x) higher at 34-36 weeks (*P*=0.028), and 0.72 log units (2.05x) higher at delivery (*P*=0.068). CRP did not differ significantly among DM-status groups in cord blood collected at delivery, the largest pairwise difference being only 0.33 log units (1.39x) between T1DM and nonDM.

**TABLE 2.**
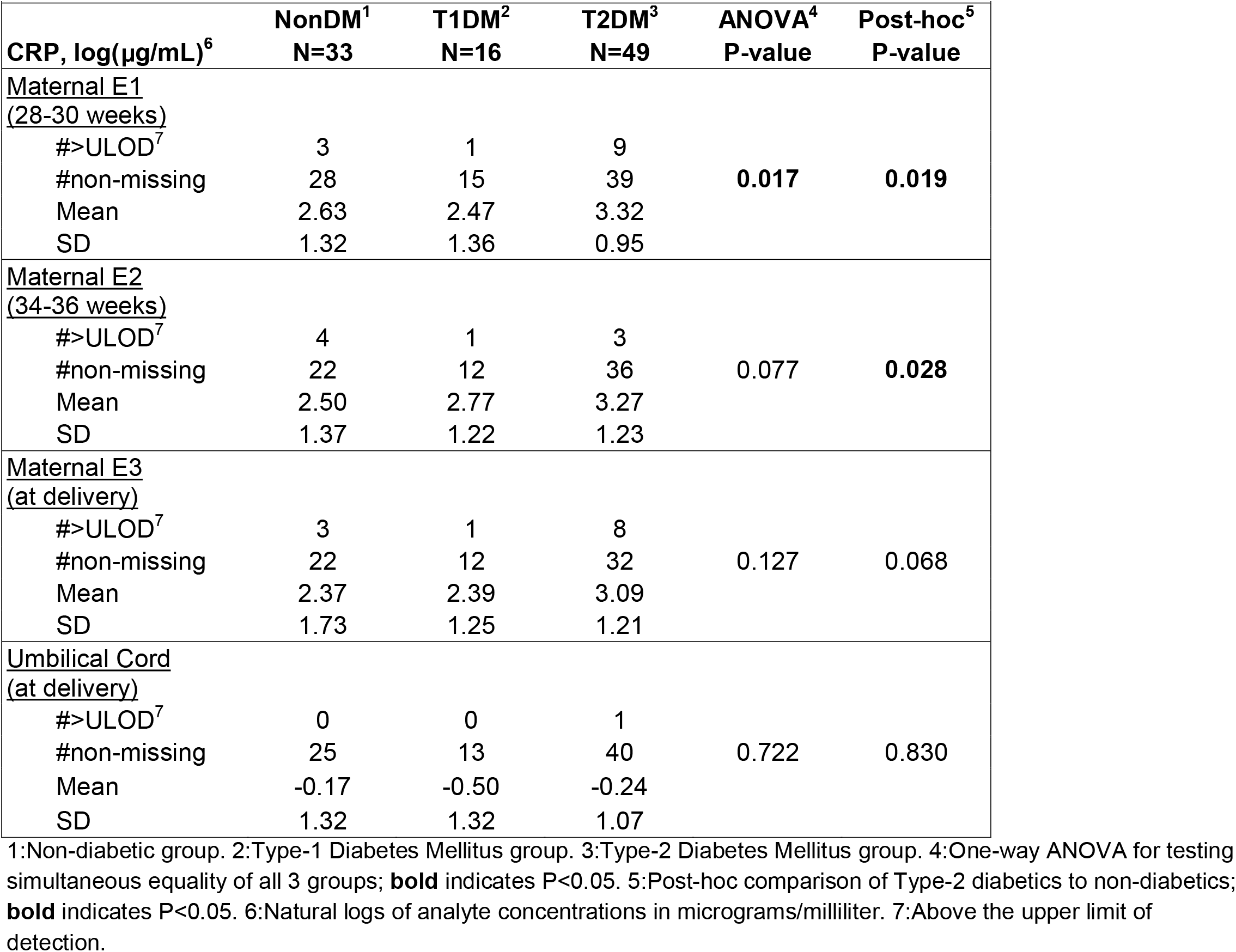
Plasma CRP levels from longitudinally collected samples.

Maternal IL-6 concentrations differed among groups in 1 of 3 time points. At 34-36 weeks (E2), IL6 levels in T1DM participants were 1.09 log units (2.97-fold or 2.97x) lower than in nonDM controls and 1.23 log units (3.42x) lower than in T2DMs (**Table 3**). Post-hoc comparisons uncovered no significant differences between control and T2DM participants. In cord-blood samples collected at delivery, the T2DM group had noticeably higher IL-6 levels than the other two groups (2.20x higher than nonDMs and 3.13x higher than T1DMs), but collectively, these differences just missed attaining statistical significance (ANOVA P=0.052).

**TABLE 3.**
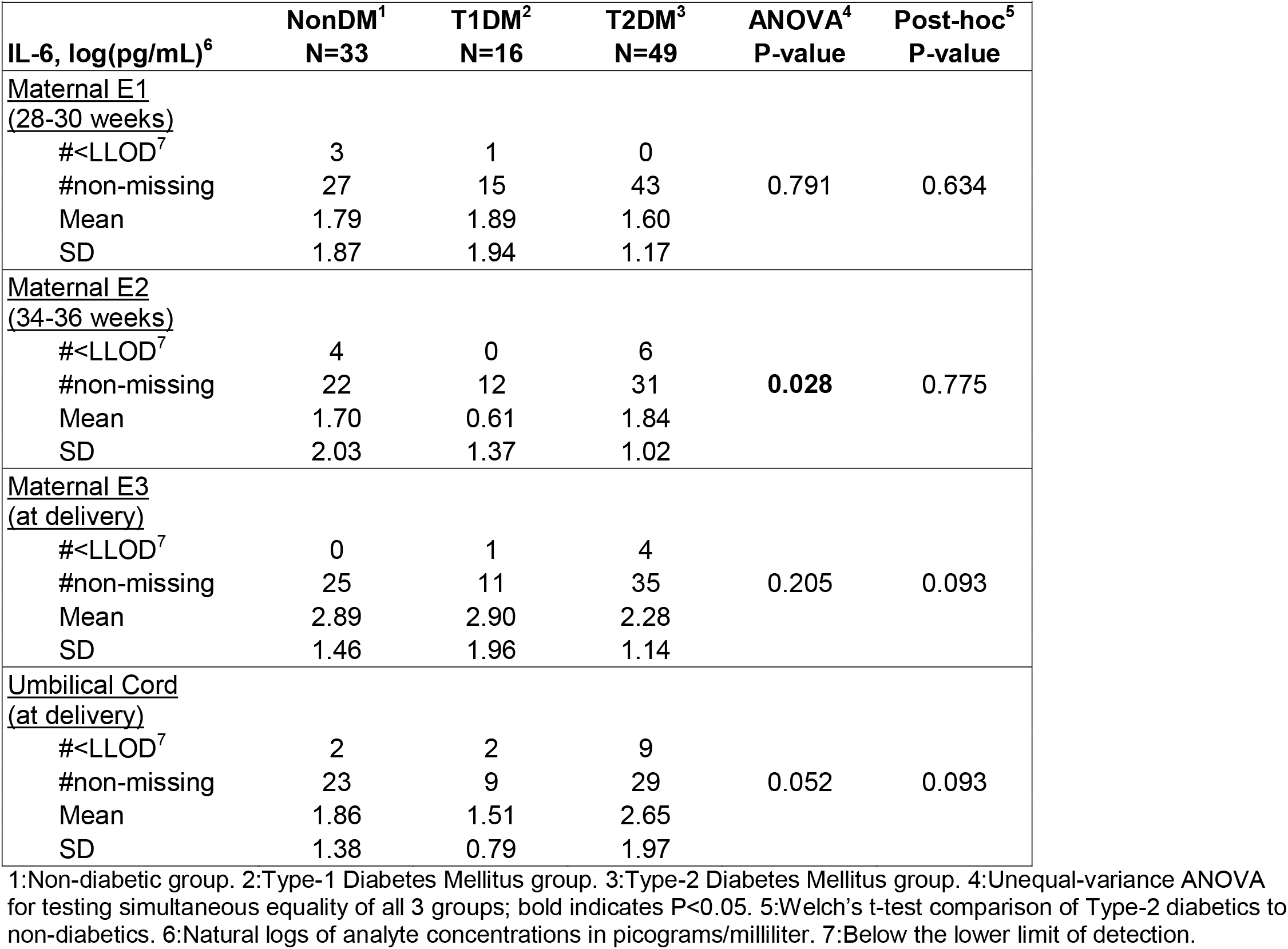
Plasma IL-6 levels from longitudinally collected samples.

Maternal BDNF concentrations were not found to be statistically different between groups in any of the maternal samples (**Table 4**). Similarly, BDNF measured in cord blood samples were not found to be statistically different between groups (**Table 4**)

**TABLE 4.**
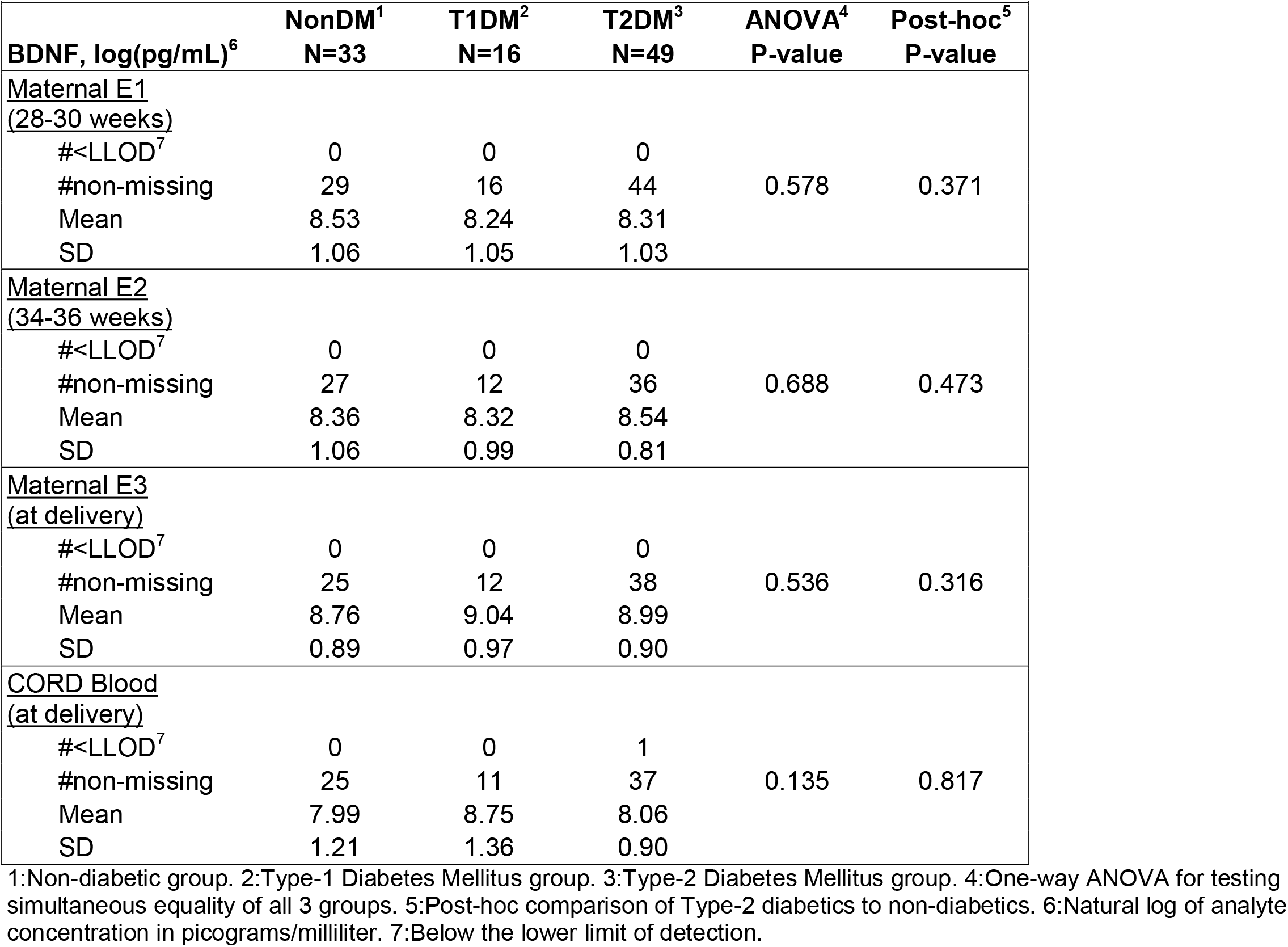
Plasma BDNF levels from longitudinally collected samples.

| TABLE 4, Plasma BDNF levels from longitudinally collected samples |  |  |  |  |  |
| --- | --- | --- | --- | --- | --- |
|  | NonDM<sup>1</sup> | T1DM<sup>2</sup> | T2DM<sup>3</sup> | ANOVA<sup>4</sup> | Post-hoc<sup>5</sup> |
| BDNF, log(pg/mL)<sup>6</sup> | N=33 | N=16 | N=49 | P-value | P-value |
| Maternal E1<br>(28-30 weeks) |  |  |  |  |  |
| #<LLOD<sup>7</sup> | 0 | 0 | 0 |  |  |
| #non-missing | 29 | 16 | 44 | 0.578 | 0.371 |
| Mean | 8.53 | 8.24 | 8.31 |  |  |
| SD | 1.06 | 1.05 | 1.03 |  |  |
| Maternal E2<br>(34-36 weeks) |  |  |  |  |  |
| #<LLOD<sup>7</sup> | 0 | 0 | 0 |  |  |
| #non-missing | 27 | 12 | 36 | 0.688 | 0.473 |
| Mean | 8.36 | 8.32 | 8.54 |  |  |
| SD | 1.06 | 0.99 | 0.81 |  |  |
| Maternal E3<br>(at delivery) |  |  |  |  |  |
| #<LLOD<sup>7</sup> | 0 | 0 | 0 |  |  |
| #non-missing | 25 | 12 | 38 | 0.536 | 0.316 |
| Mean | 8.76 | 9.04 | 8.99 |  |  |
| SD | 0.89 | 0.97 | 0.90 |  |  |
| CORD Blood<br>(at delivery) |  |  |  |  |  |
| #<LLOD<sup>7</sup> | 0 | 0 | 1 |  |  |
| #non-missing | 25 | 11 | 37 | 0.135 | 0.817 |
| Mean | 7.99 | 8.75 | 8.06 |  |  |
| SD | 1.21 | 1.36 | 0.90 |  |  |

1:Non-diabetic group. 2:Type-1 Diabetes Mellitus group. 3:Type-2 Diabetes Mellitus group. 4:One-way ANOVA for testing simultaneous equality of all 3 groups. 5:Post-hoc comparison of Type-2 diabetics to non-diabetics. 6:Natural log of analyte concentration in picograms/milliliter. 7:Below the lower limit of detection.

## 4 Discussion

The results of our study indicate that maternal diabetes, particularly type 2 diabetes, is associated with increased CRP in the third trimester of pregnancy. However, this difference did not translate to fetal CRP levels. Interestingly, IL6 was found to be lower in T1DM maternal blood as compared to both non-diabetic controls and to T2DM subjects, but only at a single time point (34-36 weeks). At two other time points in the third trimester, maternal IL6 levels were similar between groups. Fetal IL6 levels were approximately 2-fold higher in babies born to T2DM moms than non-diabetic controls but did not quite reach statistical significance. Despite no statistical differences in either maternal BDNF or cord-blood BNDF, the latter was 0.94 log-units (2.56 times) higher in neonates born to patients with type 1 diabetes compared to neonates of non-diabetic mothers.

As noted previously, increased CRP levels have been associated with FGR/SGA and schizophrenia diagnosis in offspring.^22,23^ In our study, we found elevated maternal CRP throughout the third trimester in T2DM pregnancies, potentially exposing the fetus to an environment that can influence overall fetal growth and interfere with neuronal development and function. However, despite the elevated maternal CRP, there was no elevation in fetal CRP at the time of delivery. This may indicate that CRP is not the driver of these downstream effects, but merely the marker for potential adverse outcomes. Additionally, it could be the precursor to a cytokine cascade that is the downstream cause of adverse fetal effects.

IL-6 has been associated with adverse fetal and neonatal outcomes and can be elevated in patients with diabetes.^29^ However, we unexpectedly found lower maternal IL6 in T1DM pregnant women compared to other groups at a single time point in the third trimester. It is important to note that while IL-6 has been shown to be elevated in patients with diabetes, it does not necessarily have a causal relationship with diabetes. Thus, it may be a marker for development of sequelae, but not the direct cause.^29^ Although not reaching statistical significance, we do note that fetal levels of IL6 at delivery were highest in the T2DM group. As discussed previously, offspring born to diabetic mothers can have lower IQ scores, differences in cognitive and language development, and delayed neurodevelopmental outcomes. ^4-12^ Since IL-6 has been associated with neonatal neuronal injury, IL-6 may be part of the link between these adverse neurologic outcomes and maternal diabetes. ^20^ The smaller sample size of neonatal samples compared to maternal samples may have contributed to this elevated, but non-significant finding.

In the introduction of this paper, we discussed that BDNF plays a critical part in neuronal function, synaptic plasticity and structural integrity while also having a neuroprotective role in reducing neuronal apoptosis.^25,26,30^ A study by Antonakopoulos, et al. found that fetal expression of BDNF is higher in fetuses affected by severe FGR. They hypothesized this was a response to an inflammatory condition to protect the fetal brain to limit apoptosis of neurons similar to that of early-stage neurodegenerative diseases.^31^ Despite the inflammatory nature of diabetic pregnancies, we did not find any differences in maternal or fetal BDNF between groups.

The primary strength of this study was the ability to follow a consistent cohort of patients prospectively with several data points for the majority of pregnancies. This allowed for over 200 concentrations of different biomarkers to be compared across the third trimester. Another strength of the study was the ability to match patients by gestational age. By collecting samples at predetermined gestational ages, differences in concentration due to time in pregnancy are minimized. Another strength was to match participants by BMI. Obesity is a common confounder in studies when comparing type 2 diabetes to otherwise healthy controls and our ability to match controls to diabetics helped to minimize this difference.

There are some limitations of the study, the first being the small fetal sample size. While several maternal samples were gathered across the 3rd trimester, only a single blood draw was taken on the neonatal side of the equation. However, this limitation is nearly impossible to overcome. The risk of multiple cord blood samples in otherwise healthy pregnancies is too high for a study like this, and once the neonates are born, it would be very difficult to standardize neonatal care or account for major differences in outcomes depending on gestational age at birth. Another limitation is the fact that, despite attempting to control for obesity, BMI was statistically different between groups. This was due in large part to the two following facts acting together. One, the Type-2 diabetics had an average BMI more than 9 kg/m^2^ higher than the Type-1 diabetics. Two, some non-diabetics were matched by obesity to the Type-1 diabetics whereas other non-diabetics were matched by obesity to the Type-2 diabetics. As a result, when the two subtypes of non-diabetics were combined for analysis, their average BMI was intermediate between the two types of diabetics. As obesity has been shown to influence all of the metabolites studied here, this aggregation of two different matched-control groups could have influenced results despite our matching on obesity.^32,33^ In the same vein, maternal age was a significant factor. It is possible that the length of diabetes diagnosis could have played a role in increasing serum analytes.

Future studies should work to find the definitive links between maternal inflammation and adverse fetal development. If a common pathway can be identified, targeted treatments may be able to minimize fetal effects of maternal conditions. Additionally, as our research group has found in the past, these serum analytes may influence fetal brainwave activity.^34^ As we recruit additional participants, we hope to do a more in-depth comparison of inflammatory marker concentrations to brain-wave activity using fMEG.

## 5. Conclusion

Maternal diabetes increases maternal serum markers for inflammation that have been associated with adverse fetal and neonatal outcomes persisting into early childhood. Our study confirms that maternal diabetes increases maternal CRP. More study is needed to understand the link between inflammation associated with maternal diabetes and fetal neural function and development in utero.

## Data Availability

All data produced in the present study are available upon reasonable request to the authors

## Acknowledgements

Research reported in this publication was supported by the National Institute of Child Health and Human Development under award number 1R01HD105412-01A1. Author ERS also received support from the National Center for Advancing Translational Sciences under award numbers UL1 TR003107 and UM1 TR004909. The content is solely the responsibility of the authors and does not necessarily represent the official views of the National Institutes of Health.

